# Interest in COVID-19 vaccine trials participation among young adults in China: Willingness, reasons for hesitancy, and demographic and psychosocial determinants

**DOI:** 10.1101/2020.07.13.20152678

**Authors:** Shufang Sun, Danhua Lin, Don Operario

## Abstract

**Background:** With the demand for rapid COVID-19 vaccine development and evaluation, this paper aimed to describe the prevalence and correlates of willingness to participate in COVID-19 vaccine trials among university students in China.

**Methods:** A cross-sectional survey with 1,912 Chinese university students was conducted during March and April 2020. Bivariate and multivariate analyses were performed to identify variables associated with willingness to participate.

**Results:** The majority of participants (64.01%) indicated willingness to participate in COVID-19 vaccine trials. Hesitancy over signing informed consent documents, concerns over time necessary for participating in a medical study, and perceived COVID-19 societal stigma were identified as deterrents, whereas lower socioeconomic status, female gender, perception of likely COVID-19 infection during the pandemic, and COVID-19 prosocial behaviors were facilitative factors. Further, public health mistrust and hesitancy over signing informed consent documents had a significant interactive effect on vaccine trial willingness.

**Conclusions:** High standards of ethical and scientific practice are needed in COVID-19 vaccine research, including providing potential participants full and accurate information and ensuring participation free of coercion, socioeconomic inequality, and stigma. Attending to the needs of marginalized groups and addressing psychosocial factors including stigma and public health mistrust may also be important to COVID-19 vaccine development and future uptake.

## Introduction

The novel coronavirus disease 2019 (COVID-19) pandemic has created an unprecedented global health challenge. Besides prevention, diagnosis, and treatment research, successful development and implementation of COVID-19 vaccines will be crucial to end the pandemic. Controlled human challenge trials are proposed, which involves exposing otherwise healthy individuals to the virus to test vaccine response^1^. Thus, willingness to participate in COVID-19 vaccine trials among healthy populations will be key to evaluate COVID-19 vaccines, select promising candidates, and reduce the burden of COVID-19-related mortality and morbidity. Given that vaccine trials rely on volunteers, understanding reasons for hesitancy and predictors of willingness will be important to inform ethical and scientific decisions in COVID-19 vaccine trials. Such information might also be relevant to anticipating demographic and psychosocial factors associated with future vaccine uptake, once a vaccine candidate is proven effective^2^.

The current study investigated willingness to participate in COVID-19 vaccine trials and reasons for hesitancy among young adult students in China, a population considered to be at low risk for COVID-19 mortality and with high health literacy. Demographic and psychosocial variables were explored as willingness correlates, including region of residence, gender, socioeconomic status, specific reasons for participation hesitancy (e.g., potential harm and procedural issues), and four social-cognitive variables (public health mistrust, perceived COVID-19 societal stigma, perceived COVID-19 infection likelihood, and COVID-19 prosocial behaviors). As there is no research on vaccine trials willingness regarding COVID-19, predictors were selected based on established knowledge regarding vaccine willingness and hesitancy regarding other infectious diseases (e.g., HIV, Human Papillomavirus, influenza)^3–6^.

Mistrust of health authorities has shown to affect willingness for vaccination and vaccine trials of HPV and HIV, in Europe and United States^3,5^. Mistrust could be relevant to COVID-19 vaccine trials enrollment in China due to the lack of transparency during the initial outbreak, as well as vaccine scandals in recent years^7,8^. Previous HIV vaccine research identified stigma as a barrier^9^, yet its role has not been examined in the context of COVID-19. Consistent with previous vaccine willingness research^4^, specific reasons for hesitancy, such as potential harms (physical, social) and procedural and logistical issues (e.g., consent form, time for participation), were also explored as potential deterrents. As to potential facilitators, individuals in hotspot regions (i.e., Hubei province, China’s hotspot) and from lower socioeconomic background may be particularly motivated due to their communities being heavily impacted by the pandemic^10^. COVID-19 prosocial behaviors was explored, given that altruism appears to motivate participants in other trials (e.g., HIV)^3,11^. Perceived COVID-19 infection likelihood during the pandemic may be a motivating factor due to potential indirect medical benefit in participation.

## Methods

### Participants and Recruitment

Study was approved by [masked for review]. Participants were recruited via websites targeting university students. Data was collected via an anonymous online survey, between March 20^th^ 2020 and April 10^th^ 2020, two months following the official announcement of the COVID-19 outbreak in China and during a period of state-enforced quarantine. Eligibility included (1) ≥18 years old, (2) currently enrolled in a Chinese university, (3) being fluent in Chinese. Participants were asked to read through and indicate their eligibility and consent before starting the survey. No compensation was provided. Participants were 1,992 university students. Average age was 20.38 (*SD*= 2.10, *Range*= [18, 49]). The majority were female (69.77%). Recruited participants resided in 30 provinces out of the 34 provinces of China.

### Instruments

Participants provided their demographic information (e.g., age, gender, region, socioeconomic status) and perceived COVID-19 infection likelihood, assessed by a single item: “I believe I will NOT be infected by COVID-19” (*1=completely disagree; 4=completely agree*). Responses were then categorized to perceiving infection as unlikely and likely.

### Willingness to Participate in COVID-19 Vaccine Trials

Both willingness and hesitancy items were adapted from previous research on HIV vaccine trials^4^. Willingness was assessed by a single item: “Research on COVID-19 vaccines has started, would you be willing to participate in future human COVID-19 vaccine trials when they become available?” (*1=absolutely unwilling; 2=probably unwilling; 3=probably willing; 4=absolutely willing*). Consistent with previous research^4^, willingness for COVID-19 vaccine trial participation was dichotomized (*1=willing; 0=not willing*), such that those indicted “absolutely” and “probably” willing were designated “willing” and compared against the reminder, termed as “not willing”.

Following this question, 10 items assessed **Reasons for Hesitancy** in participation. Items included a physical harm index with five items, a social harm index with two items, and three items of other concerns (full list described in Results). Participants were asked to indicate if they have each of the concerns (Yes/No/Not sure). “Yes” and “not sure” responses were treated as affirmative endorsements, and “no” was treated as absence of concerns^4^.

**Public Health Mistrust Scale**^12^ consisted of four Likert-scale items (*1=strongly agree; 4=strongly disagree*) that assessed participants’ mistrust toward the public health system in responding to an emergency (e.g., “The public health system will provide honest information to the public”). Cronbach’s *α* was 0.91.

**Perceived COVID-19 Societal Stigma** was adapted from the Perceived External Stigma of the Ebola-related Stigma Questionnaire^13^. Six Likert scale items (*1=strongly disagree; 4=strongly agree*) assessed perceived societal stigma against COVID-19 (e.g., “Most people who have had COVID-19 are rejected when others find out”). Cronbach’s *α* was 0.90.

**COVID-19-related Prosocial Behaviors** was adapted using items from two scales: the Empathic Responding to SARS scale^14^ and Prosocialness Scale^15^. Nine statements assessed prosocial behaviors specific to COVID-19 (*1=strongly disagree; 5=strongly agree*), such as donating resources, providing help to those affected by COVID-19. Cronbach’s *α* was 0.93.

### Statistical Analysis

Preliminary analysis was performed to detect univariate outliers and non-normal distributions. No outlier was detected. Bivariate analyses (Chi-square statistics, t-tests) were conducted to explore potential correlates of COVID-19 vaccine trials participation willingness. Gender (male or female), region (Hubei or non-Hubei), and socioeconomic status (low SES or not) were dummy coded. Variables that were significant at the bivariate level were entered into a logistic regression simultaneously, with willingness as the outcome variable. Consistent with prior research using the vaccine willingness scale^4^, items within the same index were collapsed into a composite score to improve the efficiency of logistic model. Adjusted odds ratio and their 95% confidence intervals were calculated.

## Results

### Willingness to participate in COVID-19 vaccine trials and reasons for hesitancy

The majority of participants (64.01%) indicated willingness to participate in COVID-19 vaccine trials (13.70% “absolutely willing”, 50.31% “probably willing”, 29.29% “probably unwilling”, and 6.70% “absolutely unwilling”). Concerns for participation were prevalent: 88.91% endorsed concerns about vaccine side effects, followed by “family may not want me to take part” (86.72%), “handicap or death from the vaccine” (84.36%), “becoming sick sooner if I ever contract COVID-19” (80.60%), “contracting COVID-19 through the vaccine” (79.86%), “time necessary to be in a medical study” (74.01%), “vaccines might contain the COVID-19 virus” (73.69%), “having to sign informed consent documents” (70.82%), “others may refuse contact with me” (65.48%), and “taking part may be seen as having COVID-19” (63.23%).

### Predictors of COVID-19 vaccine trials participation willingness

Following bivariate analysis (Table 1), variables that remained significant in predicting willingness in logistic regression were two demographic variables (lower SES, aOR = 1.49, and female, aOR = 1.27), two motivating factors including COVID-19 prosocial behaviors, aOR = 1.19, and perceived COVID-19 infection likelihood, aOR = 1.48, and three barriers including perceived COVID-19 societal stigma aOR = 0.86, hesitancy over signing informed consent, aOR = 0.55, and time necessary for a medical study, aOR = 0.60. Variance inflation factor (VIF) analysis did not suggest multicollinearity (VIF values < 3).

**Table 1.**
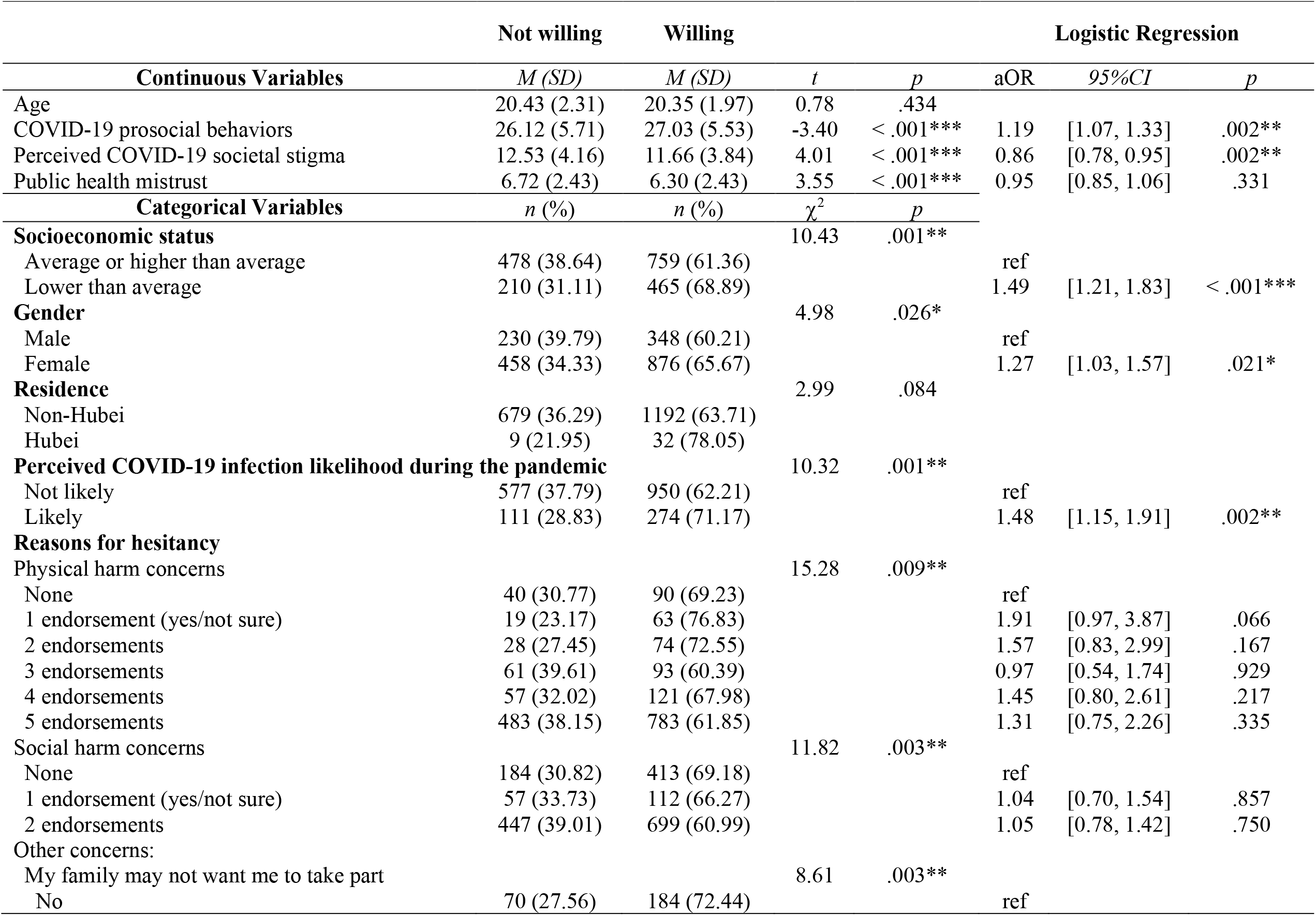
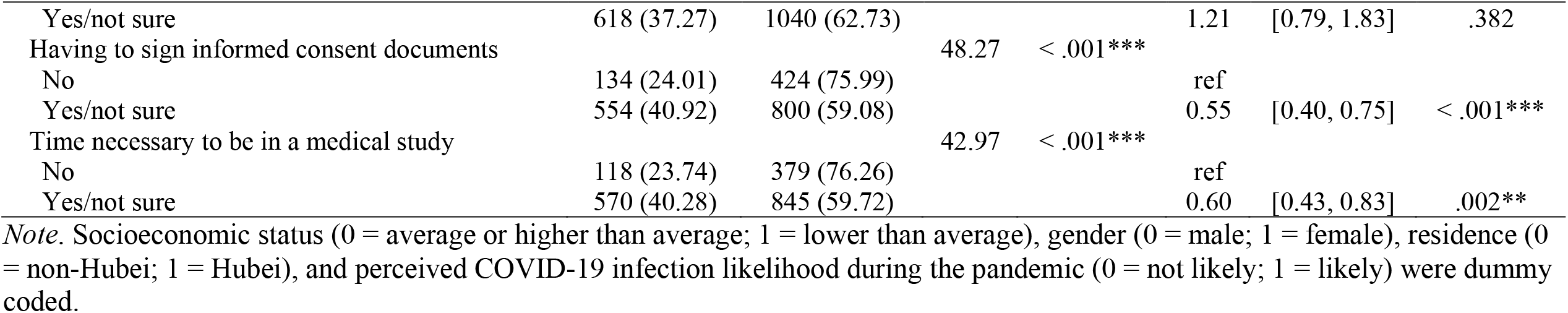
Bivariate and multivariate analysis of willingness to participate in COVID-19 vaccine trials (*N* = 1,912)

### Post-hoc analysis: The interaction of mistrust X informed consent hesitancy

In multivariate regression, hesitancy over signing informed consent documents had the largest effect size predicting willingness. We explored the role of public health system mistrust as a psychological characteristic that may interact with informed consent hesitancy to predict willingness. Accounting for all variables in the logistic regression above, the interaction of public health mistrust X informed consent hesitancy was significant, aOR = 1.29, 95%CI = [1.03, 1.62]. Main effects of mistrust (aOR = 0.78[0.63, 0.96]) and informed consent hesitancy (aOR = 0.55[0.40,0.76]) were also significant. Interaction slopes indicate that mistrust had an additive and negative effect on willingness for individuals who were not concerned about signing consent documents; for those who were concerned about signing consent documents, there was no additional effect of public health mistrust (Figure 1). Model comparison via analysis of variance revealed that the regression model including the interaction term accounted for more variance compared to the regression model without the interaction term, *p* = .028.

**Figure 1.**
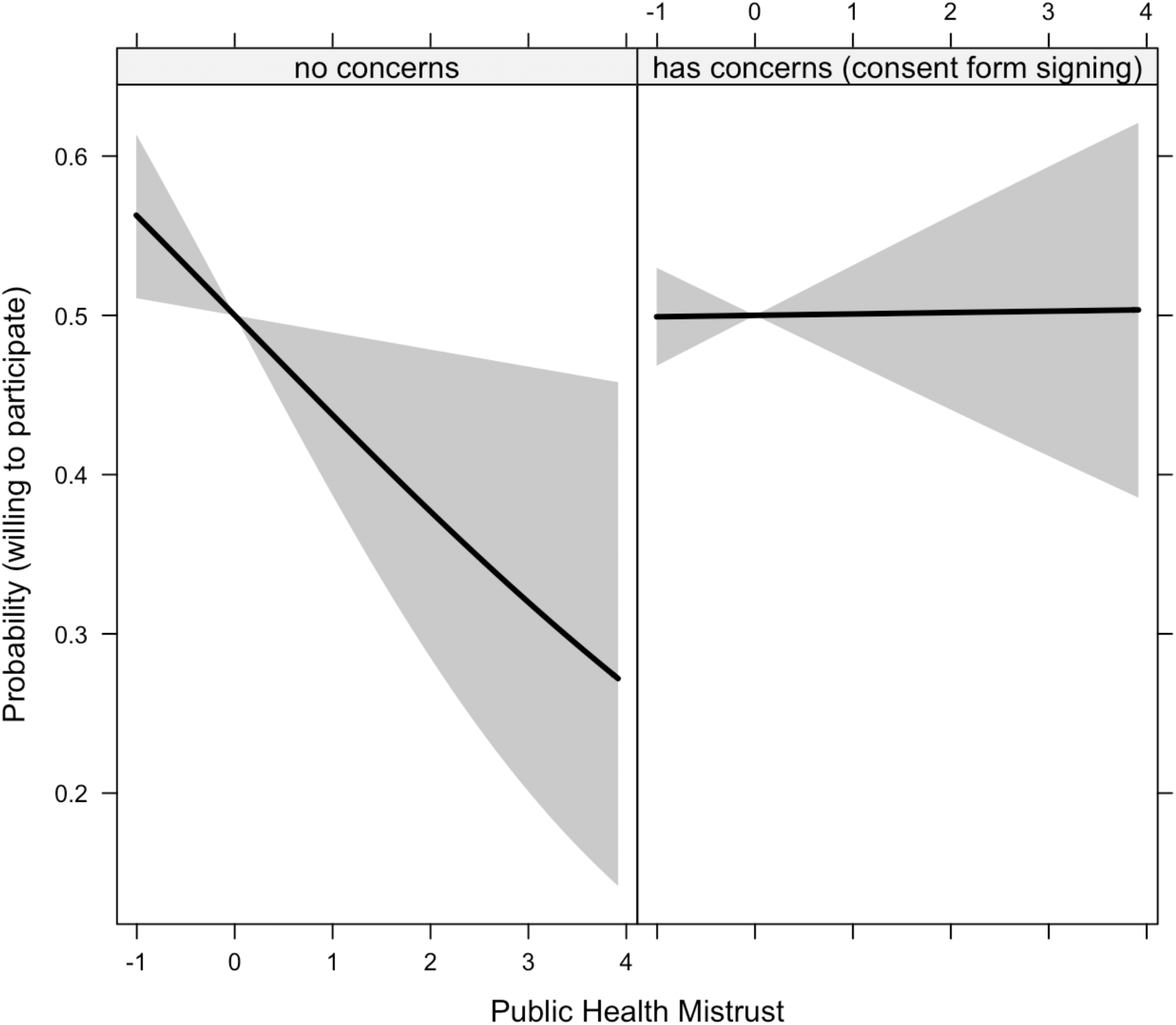
The interactive effect of public health mistrust X hesitancy over signing informed consent documents on the probability of being willing to participate in COVID-19 vaccine trials *Note*. Public health mistrust was mean-centered (*M* = 0, *SD* =1). Grey area represents 95% confidence intervals. Main effects and effects of co-variates were accounted in modeling the figure.

## Discussion

To our knowledge, this is the first study investigating COVID-19 vaccine trials participation willingness and its demographic and social-cognitive correlates. The current study found overall high willingness among young adult students in China while highlighting deterrents and facilitators important to incorporate into considerations for COVID-19 vaccine research.

Informed consent hesitancy was the strongest (and negative) predictor of willingness. Biomedical research has been growing in China, yet implementation of appropriate regulatory processes lags behind. Adverse events such as the “Golden Rice Event,” which involved deceptive language and incomplete consent content regarding risks in a health research with children, resulted in public outcries^16^. Substandard practice of consent is prevalent, including poor readability, lack of description on alternatives, and failure to provide information on procedures and rights to withdrawal^17,18^. Additionally, although designed to emphasize individual agency, some Chinese participants may view it as a transfer of responsibility for adverse consequences from researchers to participants and therefore disempowering. Thus, COVID-19 vaccine trials will need a thorough informed consent process with accessible language and adequate explanations on risks, alternatives, and rights to withdrawal to ensure rights of potential participants and research integrity. Adequate information on time required for participation will also be needed.

Public health mistrust decreased the likelihood of willingness more strongly for individuals without consent hesitancy (Figure 1). Enhancing the public’s confidence may be crucial for successful vaccine development and uptake. Such efforts may involve increasing transparency about vaccine research and its potential risks, effective regulation of vaccine production, and crisis management in situations of unexpected health emergencies.

Perceived COVID-19 societal stigma emerged as another deterrent. Given the high prevalence of COVID-19 stigma in China^19^, vaccine trials should consider potential psychosocial harm, including risks of social isolation and stigmatization due to participation. Stigma mitigation efforts will be necessary to promote willingness for COVID-19 vaccine trials and future vaccination uptake.

Facilitative factors included lower income, being female, perception of likely COVID-19 infection during the pandemic, and COVID-19 prosocial behaviors. COVID-19 vaccine trials must be careful to minimize economic coercion that might occur through incentives that inequitably drive participation among economically marginalized people. Gender-specific communication about trials participation may be needed. As participants tend to view research as therapeutic interventions^20^, those anxious about infection may be inclined to participate, perhaps due to unrealistic expectations about trial success. Similarly, young adults motivated by altruism may view societal benefits of vaccine research surpassing any concerns over personal risks. COVID-19 vaccine trials will need to facilitate potential participants to gain an accurate understanding on their infection likelihood (without the vaccine trial) and provide adequate information on risks and benefits, including a realistic depiction on the magnitude of societal benefits and influencing factors (e.g., vaccine efficacy, scale of implementation, etc.).

Study limitations include (1) lack of assessment on other factors that may contribute to participation willingness (e.g., monetary compensation, medical care provision, and vaccine administration mode); (2) cross-sectional research precluding causal inferences; (3) limited generalizability given the focus on young adults; and (4) bias due to self-report (e.g., social desirability). Study findings have implications for COVID-19 vaccine research and uptake. Rapid vaccine development has been called for, yet the high stakes and public interests involved in COVID-19 vaccine also require high standards of scientific and ethical practice. This include adequate ethical supervision, providing potential participants accurate, transparent, and accessible information about their rights, and risks and benefits associated with participation, and efforts to ensure recruitment free of coercion, socioeconomic inequality, and stigma. Public health efforts to reduce COVID-19 stigma, enhance transparency and public trust, and adequately protect marginalized communities may be critical to COVID-19 vaccine development and successful immunization implementation in the future.

## Data Availability

Data will be made available upon request.

## Acknowledgement

Work by the first author was in part supported by the Providence/Boston Center for AIDS Research (P30AI042853) and National Institute of Mental Health (T32MH078788). This research project is supported by the Fighting COVID-19 Research Fund by Beijing Normal University awarded to the second author. The funders had no role in study design, data collection and analysis, decision to publish, or preparation of the manuscript.

